# Telomere length in young patients with acute myocardial infarction without conventional risk factors: a pilot study from a South Asian population

**DOI:** 10.1101/2020.04.14.20060756

**Authors:** Mohit Gupta, Vardhmaan Jain, Manjula Miglani, M. P. Girish, Q. Pasha, Salim S. Virani, Ankur Kalra

**Affiliations:** GB Pant Institute of Post Graduate Education and Research, New Delhi, India; Cleveland Clinic Foundation, Cleveland, Ohio; Institute of Genomics and Integrative Biology, India; Michael E. DeBakey Veterans Affairs Medical Center & Baylor College Of Medicine, Houston, Texas

**Keywords:** Telomere, young, acute myocardial infarction, South Asian

## Abstract

**Background:** Myocardial infarction (MI) in the young South Asian population is rising alarmingly. There is an urgent need to identify novel markers to aid early risk estimation. Telomeres are long known to be markers of aging, and shorter telomeres have been reported in cardiovascular diseases (CVDs). Telomeres have been shown to be involved in modulating inflammation and oxidative stresses in other diseases. We aimed to identify the association of telomere length in young MI patients.

**Methods & Results:** In a case-control study of 77 subjects in each group, DNA extraction from peripheral blood leukocytes was carried out and the relative telomere length was estimated by quantitative PCR. The results were adjusted with various demographic parameters like age and gender. The correlation studies were carried out between Telomere length and lipid profile parameters (LDL, HDL, TG, TC), clinical and demographic parameters of the subjects using partial correlations.

**Results:** The relative telomere length was significantly shorter in Young MI patients (31-45 years) compared to matched healthy controls (p<0.0001). Interestingly, in a gender-based comparison, the female patients had shorter telomere length (p<0.01). Correlation analysis of the length with angiographic patterns and even the lipid profile did not reveal significance (p>0.05).

**Conclusion:** Our findings reveal the presence of shorter telomere length in patients of young MI in South Asia. Higher prevalence of stress and other factors may play a role in affecting telomere length. Larger studies are needed to confirm these findings.

## Introduction

Although of significant importance, there is scarcity of data regarding risk factors and outcomes in young individuals (< 45 years of age) suffering from acute myocardial infarction (AMI) (1). Moreover, there is growing interest in the heightened risk and etiology of coronary atherosclerosis in South Asians(2). Eukaryotic chromosomes end in tandem repeats of the deoxyribonucleic acid (DNA) sequence TTAAGGG, which are known as telomeres, and their length can be used as surrogate markers for cell aging, vascular aging, and inflammation. Accelerated telomere shortening has previously been demonstrated in patients with coronary artery disease (CAD), with recent studies supporting their causal role in CAD and other disorders (3).

## Methods

We performed a pilot study to compare telomere lengths in 77 adult (> 18 years) patients less than 45 years of age who presented with AMI, and were referred to a large tertiary care cardiovascular center in India to 77 healthy, age- and sex-matched controls without a prior history of CAD. In order to purely assess the impact of non-traditional risk factors, patients who were diabetic, had a body mass index (BMI) > 35 kg/m^2^, history of tobacco use, or were suspected of having an infectious disease or myocarditis were excluded from an original cohort of 233 patients. Genomic DNA was extracted from peripheral blood leucocytes, quantified, and assessed for telomere length by quantitative reverse transcription polymerase chain reaction (RT-qPCR) method utilizing a validated protocol (4). This technique allows measurement of relative telomere length as a ratio of telomeric repeat copy (T) DNA to a single-copy gene (S) DNA (T/S ratio). This study was approved by the institutional review board of GB Pant Institute of Post Graduate Education and Research, New Delhi, India.

## Results

In this pilot study, we found that: 1) The mean age of cases (35.33 ± 6.22) and controls (34.38 ± 5.86) was comparable (p>0.05); 2)the relative telomere length (T/S ratio) was significantly higher among controls (0.792) compared with that of cases (0.115)(P<0.05) (Figure 1a); 3) relative telomere length (T/S) was significantly higher among controls than cases in the age group 31-45 years (in the age group 18–30 years, T/S was higher among controls but didn’t attain statistical significance (Figure 1b)); 4) T/S was significantly higher among controls than cases in females (the T/S was higher in controls in males as well, but didn’t achieve statistical significance (Figure 1c)); 5) the T/S was lower in cases with anterior wall MI followed by inferior wall/posterior wall MI and lateral wall MI, although this relationship did not meet statistical significance (p >0.05).

**Figure 1:**
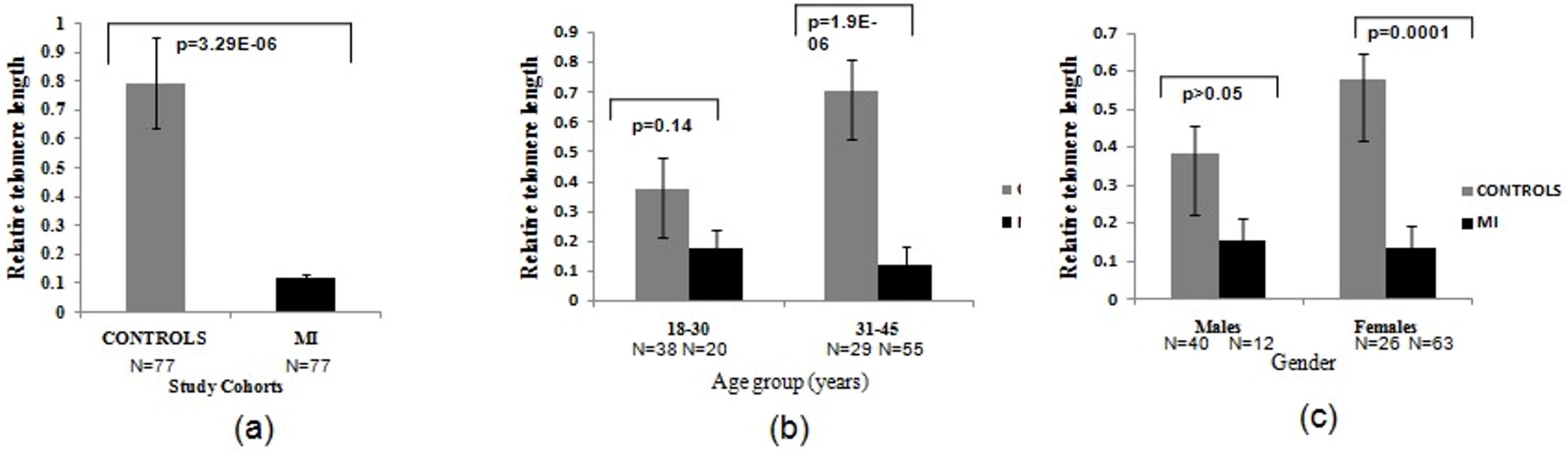
a) Relative telomere length was higher in the control as compared to Mi cases b) Telomere length was significantly higher in controls as compared to cases in the 31-45 age group, telomere length was higher in cases as compared to controls even in the 18-30 group, however statistical significance could not be achieved c) Relative Telomere length was statistically higher in female cases as compared to controls. Telomere length was also higher in male cases as compared to controls, however this relationship did not reach statistical significance.

## Discussion

To the best of our knowledge, this is the first study to compare telomere lengths in young individuals presenting with MI, who did not have other traditional risk factors for CAD. Telomeres are altered under conditions of oxidative stress, and have long been considered a marker of cell aging. Aging in itself is a risk factor for increased susceptibility for cardiovascular diseases, but chronological age may often not indicate the extent of biological aging witnessed by accelerated cellular stress—a function of genetic and environmental factors. In that sense, telomere length could be an important marker of one’s true risk for cardiovascular diseases. Moreover, atherosclerosis occurs in the setting of chronic inflammation with associated rapid white cell turnover, and consequently, may be associated with shorter telomere lengths. Thus, the extent of telomere shortening may be considered as a marker of the burden of atherosclerosis in an individual. The finding of longer telomeres in female patients compared with male patients may be attributed to the beneficial effect of estrogen which positively regulates telomerase and adds hexametric repeats to telomeres. It is consistent with the traditionally observed increased risk of CAD in males compared with females, and further strengthens the correlation of telomere length and increased cardiovascular risk. The findings of our study align well with those of a limited few that show shorter leucocyte telomere length to be associated with increased cardiovascular risk (5), but are novel in that they highlight this correlation in a young South Asian population that otherwise did not have elevated cardiovascular risk factors. There is literature to support that telomere length may be inversely correlated with modifiable risk factors including body mass index and total serum lipids (6). These findings when coupled with those of our study offer hope to explore telomere length as a tool for screening lower-risk individuals and have significant impact on public health.

The assessment of telomere length in young MI patients, especially among cases without conventional risk factors, could lead to the development of screening tools for patients at risk for CAD, and warrant further large-scale studies to confirm these findings. Although unique, our study was limited by its small sample size, its retrospective and cross-sectional nature, which do not imply causality. Moreover, adjusting for all factors that may play a role in affecting telomere length is beyond the scope of our study, and must be kept in mind while designing future studies.

## Data Availability

The authors would be willing to share patient data on a reasonable request.

